# Point-of-Care Ultrasound (POCUS) Predicts Clinical Outcomes in Patients with COVID-19

**DOI:** 10.1101/2021.04.13.21255445

**Authors:** Andre Kumar, Isabel Weng, Sally Graglia, Thomas Lew, Kavita Gandhi, Farhan Lalani, David Chia, Youyou Duanmu, Trevor Jenson, Viveta Lobo, Jeffrey Nahn, Nicholas Iverson, Molly Rosenthal, June Gordon, John Kugler, Minh Chi Tran, Xiaolin Jia, Charles Liao, Alice Cha, Evan Baum, Douglas Halket, Jai Madhok, Muhhamad Fazal

## Abstract

**Introduction:** Point-of-care ultrasound (POCUS) may detect the cardiopulmonary manifestations of COVID-19 and expediently predict patient outcomes.

**Methods:** We conducted a prospective cohort study at four medical centers from 3/2020-1/2021 to evaluate POCUS findings and clinical outcomes with COVID-19. Our inclusion criteria included adult patients hospitalized for COVID-19 who received cardiac or lung POCUS with a 12-zone protocol. Images were interpreted by two reviewers blinded to clinical outcomes. Our primary outcome was ICU admission incidence. Secondary outcomes included intubation and supplemental oxygen usage.

**Results:** N=160 patients (N=201 scans) were included. Scans were collected a median 23 hours (IQR:7-80) from emergency department triage. Triage POCUS findings associated with ICU admission included B-lines (OR 4.41 [95% CI:1.71-14.30]; p<0.01) or consolidation (OR 2.49 [95% CI:1.35-4.86]; p<0.01). B-lines were associated with intubation (OR 3.10 [95% CI:1.15-10.27]; p=0.02) and supplemental oxygen usage (OR 3.74 [95% CI:1.63-8.63; p<0.01).

Consolidations present on triage were associated with the need for oxygen at discharge (OR 2.16 [95% CI: 1.01-4.70]; p=0.047). A normal lung triage scan was protective for ICU admission (OR 0.28 [95% CI:0.09-0.75; p<0.01) or need for supplemental oxygen during the hospitalization (OR 0.26 [95% CI:0.11-0.61]; p<0.01). Triage cardiac POCUS scans were not associated with any outcomes.

**Discussion:** Lung POCUS findings detected early in the hospitalization may provide expedient risk stratification for important COVID-19 clinical outcomes, including ICU admission, intubation, or need for oxygen on discharge. A normal admission scan appears protective against adverse outcomes, which may aid in triage decisions of patients.

## Introduction

There is an urgent need to employ diagnostic modalities on the frontline of COVID-19 that are expedient, accurate, and cost-effective. Point-of-care ultrasound (POCUS) has garnered substantial interest as a potential modality to meet these needs for providers.^1–3^ POCUS devices are cheaper than traditional imaging equipment, such as X-ray or computed tomography (CT) machines, which makes POCUS ideal for surge scenarios and other resource-limited settings ^4^ Since providers using POCUS are concomitantly at the bedside assessing patients, POCUS permits an immediate and augmented evaluation of the patient, while potentially reducing personal protective equipment usage by radiology technicians and the need to decontaminate larger radiographic equipment^5^Despite the potential for POCUS to improve clinical care for COVID-19, there is limited understanding on whether POCUS can predict clinical outcomes or impending deterioration.^6,7^ In contrast, CT and X-ray findings predict important outcomes such as mechanical ventilation or death,^8–10^ which support a broader trend of using imaging modalities to provide risk stratification with COVID-19.

Previously described cardiopulmonary manifestations of COVID-19 include pulmonary edema, consolidation, pleural-line irregularities, and cardiomyopathy.^11–13^ POCUS can diagnose these pathological states with similar accuracy to computed tomography and echocardiography.^14–17^ When compared to the “gold standard” of computed tomography, POCUS has a sensitivity and specificity of 83.2% and 90.3% for alveolar-interstitial syndromes and 82.7% and 90.2% for consolidation, respectively.^18^ When compared to X-ray, POCUS is more sensitive at detecting the pulmonary manifestations of COVID-19.^19^ The sonographic manifestations of COVID-19 pneumonia include bilateral B-lines, subpleural consolidations, pleural thickening, yet with surprising absence of pleural effusions.^20–22^ These findings correlate with findings highly specific for COVID-19 observed with computed tomography (see Appendix for a full description of these sonographic findings and their CT correlates).^20,22^

There are few studies that examine the predictive utility of POCUS for COVID-19 despite recent findings that suggest imaging modalities may aid in risk stratification. In this study, we examine whether cardiopulmonary POCUS findings correlate with important clinical outcomes such as intensive care admission or need for supplemental oxygen usage, which carry substantial resource constraints. We also examine whether these findings, if detected early, are predictive of future clinical outcomes in the subsequent hospital course.

## Methods

### Participants & Setting

This was a prospective cohort study conducted at four tertiary care centers in the United States from 3/2020 – 1/2021. Our inclusion criteria included adults hospitalized with COVID-19 (based on symptomatology^23^ plus a confirmatory nasopharyngeal PCR for SARS-CoV-2) who received a POCUS examination. Patients who did not receive a POCUS scan were excluded. This study was approved by the Institutional Review Boards of Stanford University and the University of California, San Francisco. A waiver of consent was obtained by both institutions.

### Outcomes

In this exploratory analysis, previously described POCUS features for COVID-19 were compared against primary and secondary outcomes of clinical interest.^11–13^ Our primary outcome was the incidence of ICU admission vs. not among patients who received a scan. Secondary outcomes included the incidence of intubation, supplemental oxygen usage during the hospitalization or discharge, and 30-day readmission.

### Scanning Protocol and Interpretation

All patients included in this study were scanned during their initial evaluation in the emergency department or subsequent hospital days. Physician discretion determined whether to perform an initial or follow-up POCUS examination for each patient. For example, if a physician determined that a patient was clinically stable and there was no reason to perform a repeat scan, then the examination was deferred to avoid unnecessary physician exposure. Physicians were instructed to use a 12-zone scanning protocol for pulmonary views (Figure 1) and save 6 second clips of each lung zone.^24^ If a 12-zone protocol could not be performed due to the patient’s condition (e.g. the posterior lung zones were not accessible due to intubation), then a modified 8-zone protocol capturing the anterior and lateral lung fields was performed.^24^ This study utilized several POCUS devices, including Butterfly IQ™,Vave™,Lumify™, Mindray™, GE™, and Sonosite™, which represent the commercially-available portable machines at our institutions. All devices were set to the “lung” preset for lung scanning. In addition, providers acquired cardiac POCUS images using standard echocardiographic views (parasternal long axis, short axis, apical four-chamber, and subcostal) and “cardiac” presets for scanning. All collected images were saved to a local picture archiving and communication system (PACS) that was used for research purposes.

**Figure 1.**
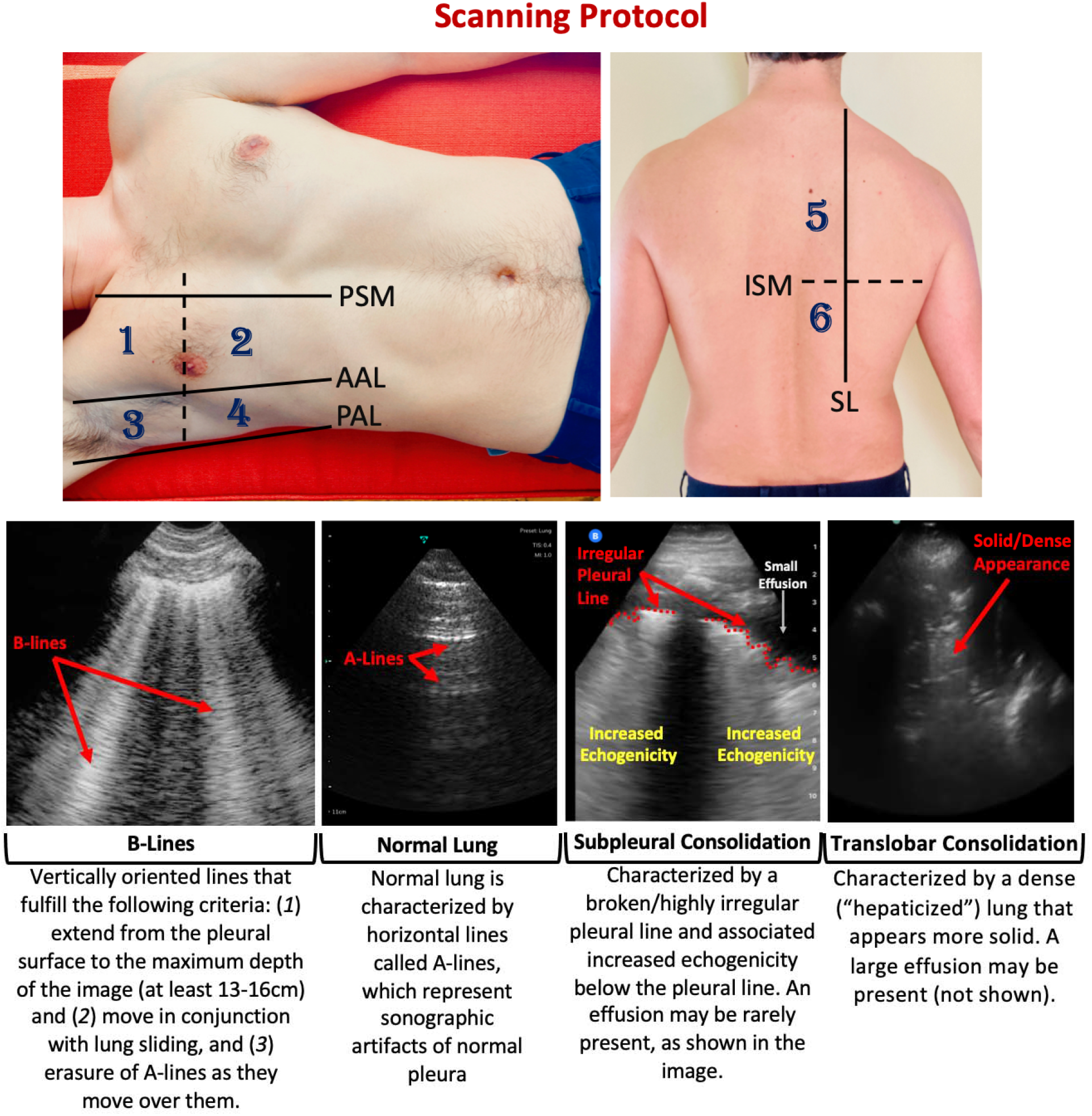
Scanning Protocol and Lung Ultrasound Findings in COVID-19 Patients. This study utilized a 12-zone protocol. On each hemithorax, there are 6 zones. The exam begins on the patient’s right side. Zones 1-2 (anterior zones) are between the parasternal margin (PSM) and the anterior axillary line (AAL) and are best obtained in the mid-clavicular line. Zones 3-4 (lateral zones) are between the anterior axillary line (AAL) and posterior axillary line (PAL) and are best obtained in the mid-axillary line. The nipple line serves as a bisecting area between these zones. Zones 5-6 (posterior zones) are medial to the scapular line (SL) and are bisected by the inferior scapular margin (ISM). The zone areas are repeated on the contralateral hemithorax (starting with zone 7). If a 12-zone protocol could not be obtained, then an eight-zone protocol (which excludes Zones 5-6) was obtained. This figure contains an overview of the observed ultrasound findings based on previously described terminology (Appendix).

The POCUS scans were obtained by physicians credentialed in POCUS for patient care at their respective institutions. The physicians involved in scanning completed a 30-minute orientation to review the scanning protocol. A core group of researchers at each site interpreted the archived images based on consensus guidelines for LUS developed by the researchers (Appendix).^24–26^ A full description of the credentials and experience of the scanners and image interpreters can be found in the Appendix. The researchers were blinded to patient outcomes when interpreting the images.

Previous investigations have demonstrated moderate to excellent interrater reliability (IRR) for LUS across different experience levels and probe types.^27–29^ Nonetheless, we conducted our own IRR analysis within the context of COVID-19 and LUS. We found that LUS has moderate-to-substantial IRR for LUS among COVID-19 for the findings included in this study.^30^ Based on our IRR findings, we developed an interpretation protocol. First, two researchers would independently apply the consensus guidelines (Appendix) to create a standardized approach to image interpretation. Next, they would input their findings into separate electronic forms. The researchers would then meet to compare their interpretations. If there was disagreement in interpretation, the two researchers would attempt to reach a consensus. If no consensus could be reached, then the data was excluded from the final database.

### Analysis

Our calculated sample size for this study was 94 patients based on reasonable assumptions (15% event rate, 80% power, alpha 0.05). Event rates were based on internal data from our hospitals at the time of study design. For the main analysis comparing ICU admission vs. not, the unit of analysis was on each scan. Therefore, the analytical set could include multiple scans for one patient that met inclusion criteria. Subgroup analysis was limited to the initial scan per patient. Chi-square and Fisher exact testing were performed for categorical variables (depending on sample size), and t-tests for continuous variables. Mann Whitney tests were performed for non-normal distributed continuous variables instead of t-tests with median and interquartile ranges (IQR) reported. Odds ratios, corresponding 95% confidence intervals and p values from the models were reported. For POCUS features with low or high rates (<5%, >95%) of events, we performed Firth logistic regressions instead to obtain more reliable estimates. Poisson regression models were performed for POCUS ultrasound count features. All analysis was performed with R statistical programming languages version 4.0.3 (Vienna, Austria).

## Results

### A. Patients & Scans

The study was sufficiently powered to analyze the primary outcome. There were N=160 patients (N=201 lung ultrasounds and N=89 cardiac ultrasounds) included in the study (Tables 1 and 2). Approximately N=32 patients received multiple LUS scans on separate days, which accounts for the greater number of LUS scans than patients. All scans were collected a median 23 hours (IQR 7-80 hours) after initial evaluation in the emergency department.

**Table 1.** Patient Demographics. Bold items denote findings of statistical significance (p<0.05). ICU, Intensive Care Unit; BMI, Body-Mass Index; COPD, Chronic Obstructive Pulmonary Disease; CVA, Cerebrovascular Accident; IQR, interquartile range.

| Characteristic | All COVID-19 Patients | Non-ICU Patients | ICU Patients | p-value |
| --- | --- | --- | --- | --- |
| **Number of Patients** | 160 | 106 (66%) | 54 (34%) | - |
| **Median Age (IQR)** | 58 (45-71) | 55 (46-69) | 60 (43-71) | 0.85 |
| **Female** | 65 (41%) | 46 (43%) | 19 (35%) | 0.45 |
| **Median BMI (IQR)** | 28.9 (25-34) | 28 (25- 34) | 31 (26-36) | 0.2 |
| **Mechanical Ventilation** | 24 (15%) | - | 24 (44%) | - |
| **Death** | 7 (4%) | - | 7 (13%) | - |
| **Median Length of Stay (IQR)** | 5 (3-12) | 4 (2-6) | 16 (9-25) | **<0.001** |
| **Supplemental Oxygen Usage** | 102 (64%) | 48 (64%) | 54 (100%) | **<0.001** |
| **Discharged on Oxygen** | 37 (42%) | 15 (27%) | 22 (65%) | **<0.001** |
| **Symptoms to Triage, Days (IQR)** | 6 [3-9] | 7.0 [3.0,10.0] | 6.0 [3.3,8.8] | 0.60 |
| **Symptoms to Scan, Days (IQR)** | 9 [5-14.5] | 9.0 [5.0,12.0] | 11.0 [5.6,17.0] | 0.08 |
| **Readmission at 30 Days** | 14 (16%) | 10 (16%) | 4 (15%) | 1.00 |
| **Medical Comorbidities** |  |  |  |  |
| **Hypertension** | 68 (43%) | 39 (37%) | 29 (54%) | 0.06 |
| **Diabetes** | 62 (39%) | 38 (36%) | 24 (44%) | 0.38 |
| **Hyperlipidemia** | 57 (36%) | 32 (30%) | 25 (46%) | 0.07 |
| **Chronic Kidney Disease** | 22 (14%) | 12 (11%) | 10 (19%) | 0.31 |
| **Coronary Artery Disease** | 20 (13%) | 15 (14%) | 5 (9%) | 0.53 |
| **Asthma** | 17 (11%) | 12 (11%) | 5 (9%) | 0.89 |
| **Current Smoker** | 17 (11%) | 11 (10%) | 3 (6%) | 1.00 |
| **History of Cancer** | 15 (9.4%) | 10 (9%) | 5 (9%) | 1.00 |
| **Congestive Heart Failure** | 15 (9%) | 11 (10%) | 4 (7%) | 0.75 |
| **COPD** | 11 (7%) | 9 (9%) | 2 (4%) | 0.42 |
| **History of CVA** | 8 (5%) | 7 (7%) | 1 (2%) | 0.36 |

**Table 2.** POCUS Findings and Primary Outcome. POCUS findings were compared between patients admitted to the ICU vs. not. On sub-analysis, scans were analyzed if they were collected within 24 hours of emergency department triage and ICU admission to examine the predictive utility of early POCUS scans (expressed as odds ratios and 95% confidence intervals). Bold items denote findings of statistical significance (p<0.05). ICU, intensive care unit; OR, odds ratio; POCUS, point-of-care ultrasound.

| Characteristic | All Patients | Non-ICU Patients | ICU Patients | P-value | OR (95% CI) From Early Scans | P-value |
| --- | --- | --- | --- | --- | --- | --- |
| No. of Scans | 201 | 132 (66%) | 69 (34%) | - | 153 | - |
| No. of 12-Zone Scans | 115 (57%) | 77 (58%) | 38 (55%) | - | 51 (33%) | - |
| Anterior Zone Scans | 198 (99%) | 130 (98%) | 68 (99%) | - | 153 (100%) | - |
| Lateral Zone Scans | 188 (94%) | 121 (92%) | 67 (97%) | - | 153 (100%) | - |
| Posterior Zone Scans | 115 (57%) | 77 (58%) | 38 (55%) | - | 51 (33%) | - |
| Normal Cardiac POCUS | 78 (39%) | 53 (40%) | 25 (36%) | 0.70 | 0.89 [0.48-1.64] | 0.72 |
| Normal Lung POCUS | 31 (15%) | 27 (20%) | 4 (6%) | **0.01** | 0.28 [0.09-0.75] | **<0.01** |
| Majority A-Line Pattern | 65 (32%) | 51 (39%) | 14 (20%) | **0.01** | 0.42 [0.21-0.81] | **<0.01** |
| B-Lines | 165 (82%) | 100 (76%) | 65 (94%) | **<0.01** | 4.41 [1.71-14.30] | **<0.01** |
| Bilateral | 124 (62%) | 71 (54%) | 53 (77%) | **<0.01** | 2.63 [1.39-5.17] | **<0.01** |
| Anterior | 121 (61%) | 68 (52%) | 53 (78%) | **<0.01** | 2.97 [1.55-5.93] | **<0.01** |
| Lateral | 132 (70%) | 82 (68%) | 50 (74%) | 0.41 | 1.33 [0.69-2.63] | 0.41 |
| Posterior | 84 (72%) | 51 (66%) | 33 (85%) | 0.06 | 2.4 [0.95-6.80] | 0.07 |
| Consolidation | 108 (54%) | 60 (46%) | 48 (70%) | **<0.01** | 2.49 [1.35-4.68] | **<0.01** |
| Translobar | 5 (3%) | 2 (2%) | 3 (6%) | 0.50 | 2.59 [0.49-16.00] | 0.26 |
| Subpleural | 100 (52%) | 59 (47%) | 41 (62%) | 0.06 | 1.68 [0.92-3.12] | 0.09 |
| Bilateral (any) | 61 (30%) | 31 (24%) | 30 (44%) | **<0.01** | 2.35 [1.26-4.41] | **<0.01** |
| Anterior (any) | 53 (54%) | 21 (39%) | 32 (71%) | **<0.01** | 3.81 [1.66-9.17] | **<0.01** |
| Lateral (any) | 69 (68%) | 32 (55%) | 37 (84%) | **<0.01** | 3.71 [1.50-10.09] | **<0.01** |
| Posterior (any) | 56 (82%) | 39 (87%) | 17 (74%) | 0.33 | 0.45 [0.13-1.57] | 0.21 |

### B. Primary Outcome: ICU Admission

Several LUS features were more common in patients who experienced ICU admission (Table 2). To assess the predictive utility of POCUS for ICU admission, we analyzed scans collected within 24 hours of ER triage or before ICU admission (n=153 scans). Several early POCUS features were again associated with ICU admission (Table 2). These included the absolute presence of B-lines (OR 4.41 [95% CI: 1.71-14.30]; p<0.01) or consolidation (OR 2.49 [95% CI 1.35-4.86]; p<0.01). The presence of either bilateral, anterior, or lateral B-lines or consolidations were similarly associated with ICU admission (Table 2). Protective factors against ICU admission included the presence of a normal lung scan (OR 0.28 [95% CI: 0.09-0.75]; p<0.01) or the presence of an A-line pattern in the majority of lung fields (OR 0.42 [95% CI: 0.21-0.81; p<0.01]. None of the patients who had an initially normal LUS within 24 hours of ED evaluation were admitted to the ICU in the subsequent 28 days.

### C. Secondary Outcomes: Intubation, Oxygen Usage, Readmission

To assess the predictive utility of POCUS on secondary outcomes, scans that were collected within 24 hours of ER triage or before ICU admission (n=153 scans) were analyzed. Early LUS findings (Table 3) associated with intubation included anterior B-lines (OR 3.10 [95% CI: 1.15-10.27]; p=0.02) and anterior consolidation (OR 6.40 [95% CI: 1.80-34.01]; p<0.01). Supplemental oxygen usage during the hospitalization was associated with B-lines at triage (OR 3.74 [95% CI: 1.63-8.63]; p<0.01), while a normal LUS at the time of triage was protective against oxygen usage for the hospitalization (OR 0.26 [95% CI: 0.11-0.61]; p<0.01).

**Table 3.** Secondary Outcomes by POCUS Findings. Scans were analyzed if they were collected within 24 hours of emergency department triage to examine the predictive utility of early POCUS scans (expressed as odds ratios and 95% confidence intervals). Bold items denote findings of statistical significance (p<0.05). ICU, intensive care unit; OR, odds ratio; POCUS, point-of-care ultrasound.

| Characteristic | Intubation OR<br>[95% CI] | p-value | Required<br>Supplemental O<sub>2</sub><br>OR [95% CI] | p-value | Required O<sub>2</sub> at<br>discharge OR<br>[95% CI] | p-value |
| --- | --- | --- | --- | --- | --- | --- |
| Normal Cardiac POCUS | 1.78 [0.76-4.18] | 0.18 | 1.36 [0.6-3.27] | 0.75 | 0.5 [0.19-1.22] | 0.13 |
| Normal Lung POCUS | 0.32 [0.03-1.32] | 0.13 | 0.26 [0.11-0.61] | **<0.01** | 0.58 [0.20-1.56] | 0.29 |
| Majority A-Line Pattern | 0.53 [0.18-1.35] | 0.19 | 0.9 [0.42-1.98] | 0.79 | 0.67 [0.29-1.52] | 0.35 |
| B-Lines | 3.82 [0.93-35.07] | 0.07 | 3.74 [1.63-8.63] | **<0.01** | 1.58 [0.61-4.35] | 0.35 |
| Bilateral | 1.01 [0.43-2.48] | 0.98 | 1.63 [0.78-3.43] | 0.19 | 1.63 [0.76-3.58] | 0.21 |
| Anterior | 3.10 [1.15-10.27] | **0.02** | 2.93 [1.39-6.34] | **<0.01** | 1.93 [0.88-4.4] | 0.10 |
| Lateral | 0.76 [0.32-1.96] | 0.56 | 2.21 [1.01-4.81] | **0.046** | 1.27 [0.56-2.95] | 0.57 |
| Posterior | 1.92 [0.53-10.32] | 0.34 | 1.43 [0.47-4.24] | 0.53 | 1.35 [0.41-4.51] | 0.62 |
| Consolidation | 2.18 [0.91-5.70] | 0.08 | 1.85 [0.88-4.05] | 0.11 | 2.16 [1.01-4.70] | **0.047** |
| Translobar | 6.38 [0.99-35.75] | 0.05 | 1.07 [0.19-10.01] | 0.95 | 2.47 [0.32-27.69] | 0.38 |
| Subpleural | 1.19 [0.51-2.89] | 0.69 | 1.67 [0.79-5.30] | 0.19 | 1.62 [0.76-3.50] | 0.21 |
| Bilateral (any) | 2.09 [0.88-4.92] | 0.09 | 1.92 [0.79-5.3] | 0.16 | 3.25 [1.35-8.11] | **<0.01** |
| Anterior (any) | 6.40 [1.80-34.01] | **<0.01** | 2.71 [0.82-9.67] | 0.10 | 2.85 [0.89-9.81] | 0.08 |
| Lateral (any) | 2.75 [0.76-14.74] | 0.13 | 2.38 [0.64-8.5] | 0.19 | 2.93 [0.83-11.66] | 0.10 |
| Posterior (any) | 0.55 [0.12-3.33] | 0.48 | 0.8 [0.07-4.95] | 0.82 | 1.53 [0.25-10.87] | 0.64 |

Consolidations present on triage scans were associated with the need for oxygen at discharge (OR 2.16 [95% CI: 1.01-4.70]; p=0.047). No POCUS findings were significantly associated with 30-day readmission, although there were several trends among specific patterns of B-lines or consolidations (Appendix).

### D. Stability of Lung Ultrasound Findings Over Time

The following analysis examined whether lung POCUS findings dynamically change over 28 days from symptom onset. Patient scans (N=201) were stratified into quartiles by time since symptom onset to their scan (Days 0-6, 7-13, 14-20, and 21-28). All scans were collected a median 9 days (IQR: 5, 14.5) from symptom onset and did not differ from patients who experienced ICU admission vs. not (Table 1). Notably, POCUS findings did not significantly change over the 28-day period (Appendix). This stability was also observed for patients who experienced ICU admission vs. not (Appendix). Similarly, there was no difference when comparing early (days 0-7) vs. late (days 21+) scans (Appendix).

## Discussion

In this prospective cohort study conducted at four medical centers, we characterize lung and cardiac POCUS findings present among patients hospitalized with COVID-19 and examine the predictive utility of these findings on clinical outcomes such as ICU admission. Several lung findings, including B-lines or consolidations, were associated with worsened outcomes such as ICU admission, intubation, or supplemental oxygen usage. Other findings, including a normal lung ultrasound or predominance of A-lines, were protective against several of these outcomes. When we confined our analysis to scans collected within 24 hours of ED triage, these findings were also predictive of clinical outcomes for the subsequent hospital course. Finally, these findings did not dynamically change over a 28-day window, suggesting that their presence, regardless of when they are detected, may be important clinical predictors.

POCUS is an expedient and cost-effective diagnostic modality that can be used in busy clinical departments to rapidly detect the cardiopulmonary manifestations of COVID-19.^5,31^ POCUS may have similar predictive utility for outcomes compared to risk stratification systems that use radiographical or laboratory markers.^8–10^ Previously described manifestations of COVID-19 pneumonia include B-lines, subpleural consolidations, and a low frequency of pleural effusions.^5,31^ These findings correlate with findings observed with COVID-19 with computed tomography (Appendix).^20,22^ Several investigators have begun to develop scoring systems to stratify patients at risk for clinical deterioration using these POCUS findings.^6,7,20,32^ Many of these scoring systems have demonstrable predictive utility,^6,7^ but they also incorporate cumbersome scanning protocols of multiple zones and may only be utilized by highly motivated users. In contrast, our findings suggest that findings of meaningful predictive utility are located primarily in the anterior or lateral lungs, which can be successfully performed by providers with more limited experience.^33^ Several of the findings (especially B-lines) have excellent interrater reliability and can be expediently learned.^30,34,35^ Based on the results of this study, a rapid assessment of the anterior lungs may provide sufficient risk stratification and warrants further study.

A challenge for providers is what to do if a scan demonstrates high-risk features, yet the patient does not require critical care. In the COVID-19 era, it is not feasible for overcrowded ICUs to accept patients who are “at risk” for deterioration. In this study, we observed that POCUS findings remained stable over the 28-day scanning period, which is consistent with previous observations for POCUS and COVID-19.^36,37^ Therefore, the detection of these findings at any time point may be concerning and warrant close observation. A potential application for POCUS in COVID-19 is for augmented risk stratification for the need for admission or discharge.^2^ Based on our results, if an otherwise stable patient presents with a normal lung ultrasound, a provider may be reassured for discharge, especially since POCUS findings for COVID-19 may remain stable over time. This practice may be supported by CT data that demonstrate pulmonary opacities often appear before symptomatology or clinical deterioration, suggesting that imaging findings can predict whether a patient will clinically worsen, even before becoming symptomatic.^38^ Further studies are needed to assess the feasibility of POCUS to guide admission or discharge decisions.

There are several limitations to this study. Certain patient conditions, such as intubation or patient mobility, prevented the provider from acquiring all 12 zones, particularly the posterior zones. Therefore, not all patients received a 12-zone scan, which limits the generalizability of the findings’ frequencies by location. Patients were scanned based on provider discretion, but it is possible that patients with certain features on X-ray or CT were more likely to receive a LUS. We did not control for patient health conditions that may have confounded the sonographic findings (e.g. interstitial lung disease), although these diseases had low prevalence in our population (Table 1). Finally, our population included only patients hospitalized for COVID-19. Therefore, our findings may not be generalizable to outpatient or triage settings, although other studies have examined the utility of POCUS in these venues.^2,7,19^

In conclusion, POCUS may provide expedient risk stratification for patients hospitalized with COVID-19, and it may have similar utility as other scoring systems that utilize radiographical or laboratory markers. Several POCUS findings, including B-lines and consolidations, were predictive of ICU admission, intubation, subsequent supplemental oxygen usage, or the need for oxygen on discharge. The predictive utility of these findings was also present when we limited our analysis to scans collected within 24 hours of admission. Importantly, a normal lung ultrasound was protective against these outcomes, even among scans collected early in the hospitalization. Future studies should determine if POCUS can be utilized to appropriate triage or discharge patients with COVID-19, especially if a simplified protocol capturing the anterior or lateral lungs is used.

## Data Availability

Data are available upon request to corresponding author

## Notes

### Competing Interest Statement

Dr. Kumar is a paid consultant for Vave Health.

### Clinical Trial

NCT04384055

### Funding Statement

No funding

### Author Declarations

Stanford University IRB and UCSF IRB provided approval for this study.

